# Feasibility and Tolerability of Deep Repetitive Transcranial Magnetic Stimulation for Mild Neurocognitive Disorder in Older Adults (Deep MIND): Study Protocol

**DOI:** 10.64898/2026.05.30.26354496

**Authors:** Mehak Ismail Rajani, Horodjei Yaya, Emily Vandehei, Anne-Marie Di Passa, Carly McIntyre-Wood, Shelby Prokop-Millar, Daniel Krzyzanowski, Molly Zhang, Allan Fein, Emily MacKillop, Jane De Jesus, Benicio Frey, James MacKillop, Dante Duarte

## Abstract

**Background:** Mild neurocognitive disorder (NCD) is a condition in which individuals experience mild cognitive decline but are independent in their activities of daily living. Due to the increasing number of people living with mild NCD and its negative impact on the quality of life, it poses a significant health burden worldwide. Thus, it warrants an urgent need for innovative approaches to address the lack of effective treatment options. Deep transcranial magnetic stimulation (dTMS), a non-invasive neuromodulation technique approved for the treatment of various neuropsychiatric disorders, could serve as a novel intervention for mild NCD. It can stimulate deeper and broader areas of the brain implicated in mild NCD, such as the prefrontal cortex, insula, and anterior cingulate cortex.

**Objectives:** This study will examine the feasibility and tolerability of the Health Canada and Food and Drug Administration (FDA) approved dTMS coils (H1, H4 and H7 coils) in individuals with mild NCD. Secondarily, it will assess the impact of dTMS on cognition, mood, sleep, anxiety, brain activity (via electroencephalography), and blood biomarkers of neurodegeneration and inflammation.

**Methods:** This open-label pilot study will recruit a total of N=30 participants between the ages of 60-90 with mild NCD. Participants will be assigned to one of the three dTMS coil conditions (H1, H4 & H7) and will complete a total of 20 dTMS sessions over 6 weeks. Data will be collected before, during, immediately after, and one-month following the intervention period.

**Discussion:** This pilot study will generate necessary evidence regarding the feasibility and tolerability of dTMS in mild NCD. This will be used to determine whether a definitive trial is justified and inform the trial procedures. In the long term, dTMS may address a critical gap in therapeutic options for mild NCD.

**Clinical Trial registration:** The protocol was registered on Clinicaltrials.gov (CT07038798) on June 2^nd^, 2025.

## Background

Mild neurocognitive disorder (NCD) is characterized by the onset of mild cognitive decline in one or more cognitive domains that does not interfere with one’s ability to perform activities of daily living (1). The diagnostic criteria for mild NCD share substantial overlap with mild cognitive impairment (MCI), a related clinical construct initially conceptualized as an at risk or transitional state between normal cognitive aging and dementia (2). Based on a recent meta-analysis, 23.7% of older adults are affected by mild NCD worldwide (3), with an increased prevalence in older age groups (6.7% at 60-64 years vs 25.2% at 80-84 years). These numbers are projected to rise due to increased longevity and a rapid increase in the global geriatric population, which is estimated to be 2.1 billion by 2050 (4).

Furthermore, this upward trend in the number of people affected with mild NCD is complicated by the high conversion rate of mild NCD to dementia, with a rate of up to 10-15% in high-income countries, to as high as 15.4-33.4% in low-income countries(5). Additionally, mild NCD reduces quality of life, with a high proportion of patients reporting social isolation and lower self-esteem following diagnosis attributed to partial dependence in performing complex tasks (6). Collectively, this leads to increased healthcare costs and economic burden (7).

Despite the high burden at both the individual and societal levels, there is no current gold-standard treatment available for mild NCD. The existing treatment interventions, namely, pharmacological, cognitive training, and lifestyle modifications, are mainly aimed at reducing the risk of progression to dementia, but do not cure the symptoms (8). This underscores the pressing need for new treatment strategies for mild NCD.

Mild NCD are a heterogeneous group of disorders as the etiology can be attributed to a myriad of underlying neurological mechanisms, including impaired functional connectivity, reduced brain volume, activity and synaptic plasticity, and high levels of inflammation (9). Functional MRI studies report disturbed functional connectivity within large-scale networks, including the default mode network (DMN) and the salience network (SN) (10), as well as reduced functional connectivity involving the bilateral dorsolateral prefrontal cortex (DLPFC) and parietal lobe and sub-cortical regions compared to healthy controls (10). This disconnection is compensated by increased connectivity between the left DLPFC and the right prefrontal cortex especially during cognitive tasks, compared to healthy older adults (12). Thus, targeted treatments to improve connectivity between neurons and plasticity in the concerned brain regions offer a promising avenue to enhance cognition in individuals with mild NCD.

Recently, non-invasive neuromodulation techniques, such as transcranial direct current stimulation (tDCS) and repetitive transcranial magnetic stimulation (rTMS), have shown promising results in improving executive functions and memory, respectively (9). These treatments work by depolarizing neurons using electrical stimulation (tDCS) or magnetic pulses (rTMS) of varying intensities to either excite or inhibit neuronal function. However, both techniques are superficial, and the field of strength decreases with depth (13). A relatively newer technique called deep transcranial magnetic stimulation (dTMS), operating on similar principles as rTMS, penetrates broader and deeper into the brain, specifically up to 6 cm in comparison to a more superficial field depth in rTMS (1-4 cm)(14). This is achieved by using a unique electromagnetic coil called the “Hesed-coil” or “H-coil”(14). Like rTMS, dTMS delivers repetitive electromagnetic pulses into the brain to generate an electric current; however dTMS maintains a higher electric field amplitude at greater depths (70-80%) in comparison to the traditional rTMS figure-8 coil (50%) (14). This is especially critical for our study population, as evidence illustrates that older individuals may require deeper stimulation to account for cortical atrophy (15,16).

Three H-coils (H1-, H4- and H7-) have been approved by the U.S Food and Drug Administration (FDA) and Health Canada for several psychiatric disorders, with each targeting specific brain regions as determined by their physical configuration. The H1-coil was approved for the treatment of treatment-resistant depression in 2013, by stimulating bilateral prefrontal cortices with a preference over the left hemisphere (17). The same region, particularly the left DLPFC, is vital for inductive reasoning and has diminished activation in mild NCD (18). Similarly, the H4-coil, which received clearance for smoking cessation in 2020, stimulates the bilateral insula and prefrontal cortices. The insula is important for social cognition(19) and shows abnormal functional connectivity in mild NCD (20). Lastly, the H7-coil, which received marketing authorization for the treatment of obsessive-compulsive disorder (OCD) in 2018 (21), and anxious depression in 2022 (22), engages the anterior cingulate cortex (ACC) and medial prefrontal cortex. These regions exhibit impaired neural activity in individuals with mild NCD as observed by slower information processing and suboptimal task execution on prefrontal event-related potentials (ERPs) (23). Additionally, these regions are part of DMN and/or SN, which also show disrupted functional connectivity in mild NCD (24).

Although there is strong rationale for the hypothesis that dTMS would be beneficial for cognitive functions (25,26), its effect on mild NCD remains under-researched. To date, only one pilot study has examined the combined effects of cognitive training and dTMS in people with mild NCD using the H7-coil (27). This pilot trial reported significant improvement in the memory domain maintained for up to six months. Nevertheless, evaluating dTMS as a standalone therapy for mild NCD requires further research. Given the clear need for additional research in this area, our study aims to assess the feasibility and tolerability of the H1-, H4- and H7-coils in older adults with mild NCD. The secondary aims include to (i) measure changes in cognitive status using validated, subjective and objective neuropsychological tools and other clinical symptoms, such as mood, anxiety, and sleep quality; and to (ii) identify potential blood and electrophysiological biomarkers using plasma analyses and electroencephalographic (EEG), respectively.

To achieve greater insights into the impact of dTMS in individuals with mild NCD, we will analyze EEG recordings of participants before, during, and after dTMS treatments. Evidence suggests that different forms of dementia are associated with unique electrophysiological markers. For example, Jelic & Kowalski (2009) (28) reported increased theta power, decreased alpha power, and decreased alpha and gamma coherence in patients with progressive MCI. Additionally, a cross-sectional study (29) reported the anteroposterior localization of alpha power shows promise as a biomarker for NCD progression in people with Alzheimer’s disease. Given that EEG not only provides a distinct profile associated with mild NCD diagnoses but also reveals predictors of progression to more advanced disease, the findings from our study will help elucidate the neurophysiological underpinnings of mild NCD, identify individuals at risk for disease progression, and contribute to the growing body of literature on precision medicine.

## Methods

### Study Design

This is a prospective, longitudinal, one-way, three-group, within-subjects, and open-label pilot study. This is designed to assess the feasibility and tolerability of dTMS treatment in patients with mild NCD. Each participant will receive 20 dTMS treatments over a period of 6 weeks and will be followed up after a month, making it approximately a 10 week-long commitment. The total duration of the study is estimated to be approximately 24 months (see Figure 1). The study protocol received approval from the local Hamilton Integrated Research Ethics Board (HiREB) (Project ID: 18440) and is registered on Clinicaltrials.gov (CT07038798).

**Figure 1.**
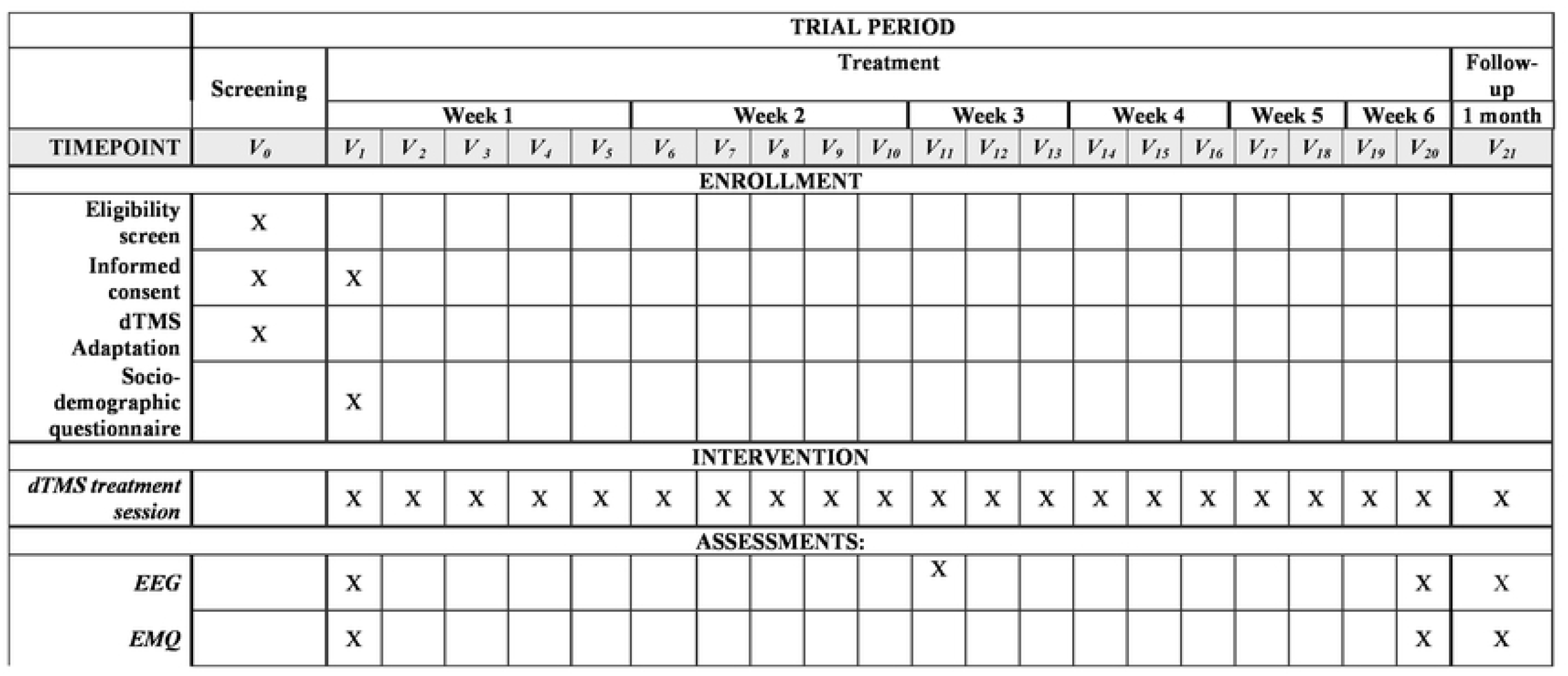
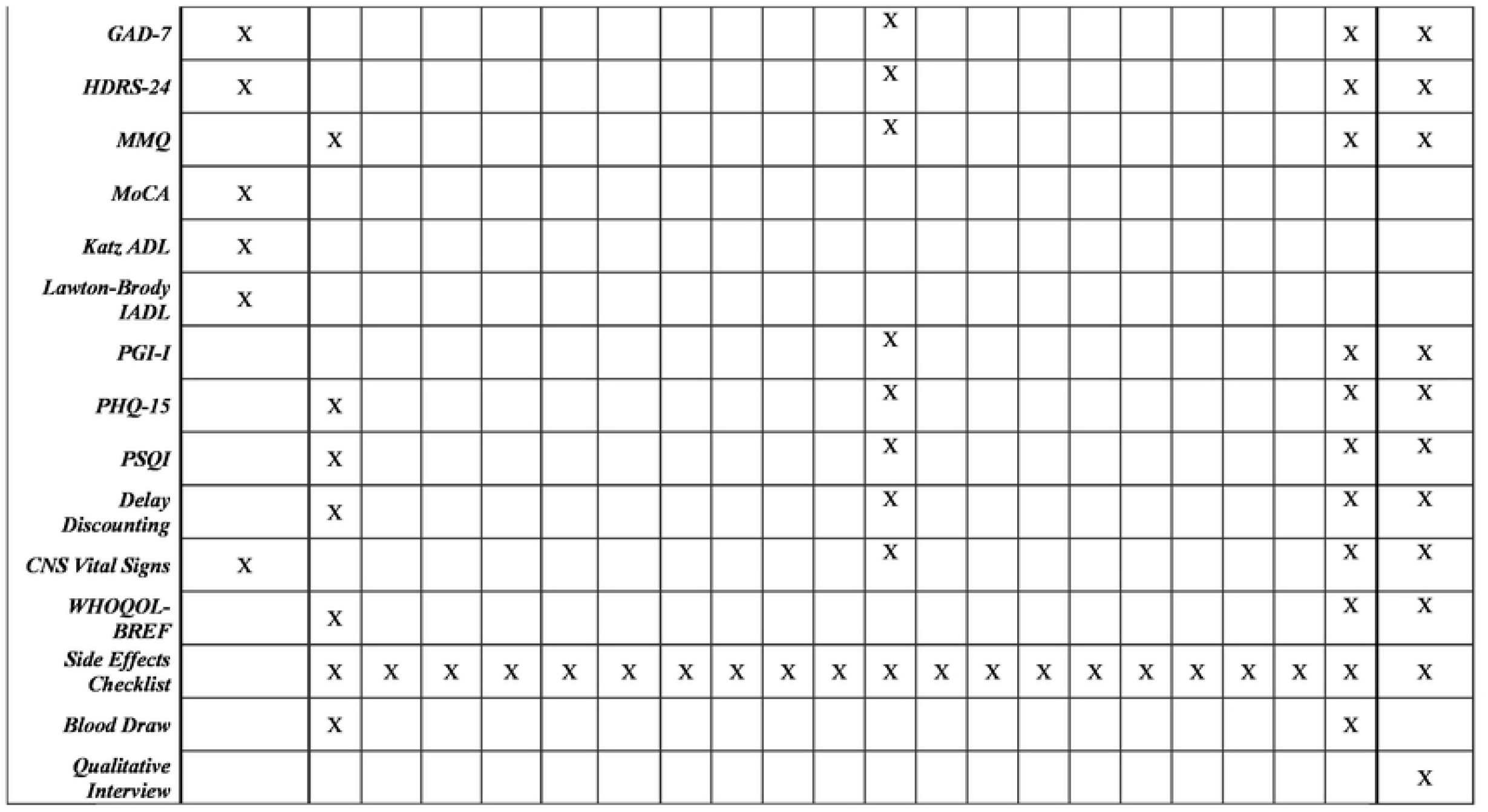
Schedule of the study enrollment, assessments and interventions. *V*_0:_ visit 0. screening of the study participants: *V*_*1*_.*V*_*21:*_: visits 1-21: **<u>Abbreviations:</u>** EEG=electroencephalogram; EMQ = Everyday Memory Questionnaire; FU= follow up; GAD-7 = generalized anxiety disorder-7 item; HDRS-24_c_ Hamilton Depression Rating Scale 24-itcm; KatzAOL_c_ Katz Index or Independence in Activities of Daily Living; Lawton-Brody IAOL = Lawton-Brody Instrumental Activities of Daily Living scale; MMQ = Multifactorial Memory Questionnaire; MoCA = Montreal Cognitive Assessment; PHQ-15: Patient Health Questionnaire Physical Symptoms; PGl-1 = Patient Global Impression - Improvement; PSQI = Pittsburgh Sleeping Quality Index; SE Checklist= Side Effects Checklist; WHOQOL-BREF = WHO Quality of Life-BREF. *Note*. Assessments and questionnaires may be conducted+/-1 day to allow for flexibility. Although the study aims 10 accomplish 20 stimulation sessions, we will accept a minimum of 15 stimulation sessions (i.e., ≥ 75% of intended sessions). We will offer to compensate for missed sessions at the end of the stimulation series.

### Eligibility Criteria, Participant Characteristics, and Setting

The study is currently underway at the Peter Boris Centre for Addictions Research (PBCAR) at St. Joseph’s Healthcare Hamilton (SJHH), Ontario, Canada. A total of 30 individuals between the ages of 60 90 years with mild NCD will be enrolled in this study. We will employ a sex equity approach, allowing for a maximum imbalance of 70/30% of female and male participants from either side, per arm. The investigation will enroll 10 older adults per coil (H1-, H4-, H7-).

The inclusion criteria are as follows: (i) 60 - 90 years old; (ii) able to provide informed consent; (iii) subjective concern of mild decline in cognitive function over the past year; (iv) mild impairment in cognitive performance as evidenced by a score between 18-26 on the Montreal Cognitive Assessment (MoCA); (v) preserved independence in everyday activities (a score of 8/8 on the Lawton-Brody Instrumental Activities of Daily Living scale); (vi) independently mobile (a score of 6/6 on Katz Index of Independence in Activities of Daily Living); and (vii) stable dosages of other psychotropic medications for at least 4 weeks prior to screening.

Exclusion criteria include: (i) currently receiving treatment or have a subjective need for treatment for bipolar I disorder, bipolar II disorder, or a psychotic disorder; (ii) active suicidal behavior; (iii) severe depression and/or anxiety (characterized by a score of >19 on the Patient Health Questionnaire Physical Symptoms-9 (PHQ-9) or >24 on the HDRS24, and/or a score of >14 on the GAD-7); (iv) other neurological or psychiatric disorders accounting for the cognitive deficits; (v) impairment in basic and/or instrumental activities of daily living; (vi) substance use disorder (other than tobacco use disorder) in the past three months before entering the study; (vii) traditional contraindications to rTMS: intracranial or metal implants in the head or nearby regions, excluding the mouth, that cannot be safely removed; history of epilepsy or seizures; active unstable medical condition (recent laboratory and neuroimaging alterations, delirium); pacemaker and/or implantable cardioverter-defibrillators; current use of bupropion (30), treatment with equivalent benzodiazepine dose to lorazepam >2 mg/day (31,32); (viii) severe literacy, visual, or hearing issues that affect the ability to engage in the interviews. (ix) recurring migraines or headaches (weekly or more); (x) frequent dizziness/vertigo; (xi) individuals residing beyond the borders of the Greater Hamilton Area and its neighboring vicinities.

### Sample size

As a pilot and feasibility study, we did not formally perform sample size calculation, however, based on previous feasibility trials on the use of TMS we project that this will provide us enough data to leverage on for a larger clinical trial.

### Recruitment

The study is being advertised using a multi-modal approach: digital outreach, community-based, and clinical site-based recruitment. To reach our specific patient demographics, email promotions have been sent out using targeted mailing lists from the Seniors Mental Health Service at SJHH, study information is being displayed at local hospital campuses, awareness talks are being given at senior’s mental health and cognitive remediation programs (e.g., Learning the Ropes for People Living with MCI), and geriatric clinics have been engaged to identify and refer appropriate patients to the study. To reach a broader audience, study information has been posted at local libraries, community centers, seniors clubs, recreation centers, and retirement homes. Additionally, traditional forms of advertisement such as social media (i.e. Meta platforms), transit, and newspaper advertisements, are also being utilized. Recruitment for this trial started in June 2025 and will run for 18-20 months. As of May 12, 2026, 203 individuals completed the online-screening form, out of which 87 who were possibly eligible, were contacted. 32 accepted the invitation for in-person screening, and 9 participants have been enrolled for the H1-coil.

### Screening

Participants are being screened through a two-stage screening process. In stage one, prospective participants complete an online screening form obtained through our various advertising platforms. In stage two, individuals who meet the pre-screen eligibility criteria undergo a comprehensive in-person screening, during which research personnel collect detailed medical history and administer screening assessments. Written consent will be obtained at both stages of the screening (See Figure 1).

### Primary Outcomes: Feasibility and Tolerability Measures

The primary aim of this study is to establish evidence for the tolerability and feasibility of dTMS in older adults with mild NCD. For tolerability, patients will be assessed for adverse events and if these events contribute to study withdrawal. Furthermore, feasibility will be measured by protocol completion rate (i.e. % of dTMS sessions completed); retention rate (i.e. % of participants who completed the study after enrollment); screening rates and capacity (i.e. n screened; n enrolled as a percentage of n screened monthly); recruitment rate and capacity (i.e., n per month and total n enrolled monthly); and duration of intervention and assessment processes. Lastly, we will conduct qualitative interviews to explore the effects of dTMS not captured by the quantitative methods and explore if dTMS is an acceptable form of treatment.

### Secondary Outcomes: Impact on Clinical Outcomes

Participants will be assessed for cognitive performance, depressive and anxiety symptoms, sleep quality, and somatic symptoms through standardized tools. See Figure 1 for the schedule of assessments and tools used following the standard protocol items: recommendations for interventional Trials (SPIRIT) guidelines and checklist (see S1 for SPIRIT Checklist and S2 Appendix) for description of the protocol (33).

### Biomarkers

#### EEG

To record dTMS-induced changes in the cortical excitability, resting state EEG of the participants will be collected with eyes open and closed for 6 minutes each at baseline, midpoint (visit 11), endpoint (visit 20) and at one-month follow-up.

#### Plasma

We will measure specific biomarkers associated with neurodegeneration and pro- and anti-inflammatory cytokines, which include: brain-derived neurotrophic factor (BDNF), beta-amyloid ratio (Aβ1-42/Aβ1-40 ratio), phosphorylated Tau 217 (pTAU-217), neurofilament light chain (NfL), and IGF-1, VEGF, TGF-β1, MCP-1, IL-18). These will be measured at baseline and at visit 20 to assess the effect of dTMS treatment on biomarkers levels. The sample will be collected by trained phlebotomists at the Hamilton Regional Laboratory Medicine Program Specimen Collection Centre at SJHH West 5th campus. Samples will be stored, processed, and analyzed at the Centre for Clinical Neurosciences (SJHH West 5th). Serum samples will be analyzed using the following methods: enzyme-linked immunosorbent assay (ELISA) and single molecular array (SIMOA) technology.

### Intervention

dTMS will be administered using the BrainsWay dTMS device (BrainsWay™ Ltd. Jerusalem, Israel) using the H1-, H4- and H7-coils. Only certified research team members will administer the treatment.

At in-person screening, resting motor threshold (RMT) will be assessed to determine the minimum stimulation intensity required to elicit a motor response on the abductor pollicis brevis muscle. This will be determined using both observation and electromyography. This will be followed by a 15-minute exposure to dTMS. Individuals who are still interested in participating will then be enrolled in the study.

For treatment, the helmet will be positioned, at 6 cm for H1- and H4-coils, and 4 cm for H7-coil anterior to the RMT spot on the scalp. All dTMS treatments irrespective of the coil, will be administered at 18 Hz frequency, consisting of 55 trains, with a 2-s train followed by 20-s rest, for a total of 1980 pulses. Each dTMS session will be of 20 minutes. During the initial treatment phase, the treatment intensity (% of RMT) will be increased gradually to enhance tolerability to a maximum of 120% of RMT.

The participants will undergo 20 dTMS sessions over a 6-week period. The initial 10 dTMS sessions will occur daily (five per week), followed by 3 sessions per week for the following 2 weeks and finally, tapering off with 2 sessions per week for the last two weeks. Eligible participants will be assigned to the coils sequentially based on the enrollment time. The first 10 participants will be assigned to H1, the next 10 to H4, and the final 10 to H7-coil. The assignment will be non-randomized and the investigators will be aware of the coil assignments.

### Data Management and Availability

Data collection began in June 2025 and is expected to be completed within 18-20 months from the start of the trial. All data will be collected and stored using electronic data capture systems (i.e., REDCap) and case report forms (CRFs). To ensure confidentiality, identifiable data will be segregated from coded data and stored in a locked cabinet within a locked office at PBCAR and on a password-protected folder in a shared drive that can only be accessed by the research team members. Upon completion of data collection, the data will be cleaned, analyzed and disseminated through various conference presentations and publications. This process will take approximately 4 months to complete. Upon publication of the results, the confidentiality of the participants will be maintained, and none of the personal identifying data will be released or published without consent. Data and study materials will be subject to HiREB audits or legally mandated requests. All participants are informed of these possibilities at the time of signing the consent form.

### dTMS Safety and Risk Management

dTMS is generally highly tolerable as established in younger populations. With respect to older adults, our team was able to establish evidence of high tolerability for dTMS (100% of participants received a therapeutic dose) in a recent pilot study in older individuals with depression (34). Common side effects of dTMS include tension headaches and scalp and neck pain, mainly due to the sensory stimulation and the pressure exerted by the coil. These side effects are often resolved with over-the-counter pain medications (12). However, a rare serious side effect associated with dTMS is the risk of seizure in approximately 6 per 10,000 patients (35). Although the risk of seizure is extremely low if correct procedures are followed, through our two-step screening, our team will thoroughly assess for contra-indications for dTMS. This process will identify individuals with a high risk of seizure. Participants are informed of all risks associated with dTMS within the consent form.

### Statistical Analyses

Normality and homogeneity of variances for all continuous variables will be assessed using the Shapiro-Wilk and Bartlett tests, respectively. Descriptive statistics will be reported as either mean± standard deviation (SD) for parametric or median interquartile range (IQR) for non-parametric variables. Missing data will be handled using multiple imputation, assuming that they are missing at random. Comparisons of categorical variables, including sex, protocol completion rate, retention rate, screening rate, recruitment rate, safety and tolerability, between conditions will be made. For continuous variables, the Wilcoxon-Mann-Whitney non-parametric test will be used due to small sample size. We will use the data generated from secondary aims to compare pre- and post-treatment scores descriptively and conduct mixed analyses of variance (ANOVA). Furthermore, EEG data, including alpha, theta, and gamma spectral power, connectivity, and coherence in fronto-temporo-parietal regions will be analyzed using standardized data processing pipelines in Python and MATLAB. We will also study cross-frequency coupling (theta (4– 8Hz)–gamma (>25Hz) phase–amplitude coupling) and phase synchronization in the upper theta frequency band between prefrontal and temporal areas. Changes in cognitive performance and other clinical outcomes will be correlated with the changes in these EEG metrics. Lastly, qualitative outcomes will be analyzed using thematic analysis via NVivo software.

## Discussion

This study will primarily investigate the tolerability and feasibility of dTMS in individuals with mild NCD. We will examine protocol completion, recruitment and retention rates, and patient acceptability of this modern treatment approach for mild NCD. Additionally, we will assess if there are any preliminary benefits of dTMS treatment in managing clinical symptoms associated with mild NCD. Furthermore, this study will also track plasma proteins of neurodegeneration and inflammation, and EEG, which may help quantify treatment response and identify predictive biomarkers.

This pilot study has a number of strengths: (i) while a previous study evaluated the impact of dTMS in combination with cognitive training on mild NCD, the current study, to our knowledge, will be the first to investigate the effects of dTMS as a mono-therapeutic intervention for mild NCD; (ii) the outcomes of this study will support efforts to establish a full-scale randomized clinical trial (RCT) investigating dTMS as a potential treatment for mild NCD in older adults; (iii) any barriers in the study design (for example number of visits, treatment compliance, recruitment strategies, retention rates etc.) can be addressed in the design of the definitive RCT; (iv) data obtained from this pilot study will serve as the basis for a power analysis estimating sample size for the future RCT; (v) while this pilot study is essentially gathering data regarding the feasibility and tolerability of dTMS in older adults, the participants may benefit from the clinical impact of dTMS on cognition; (vi) this study may help mitigate the social stigma associated with a diagnosis of mild NCD as the participants will regularly interact with the study staff and openly discuss their cognitive challenges in a supportive atmosphere; (vii) the study participants will gain awareness about mild NCD, receive information about community resources, and if they are not already seeking professional help for cognitive concerns, obtain a recommendation for referral to a specialist for ongoing care; (viii) lastly, this study may also identify potential plasma and electrophysiological biomarkers of treatment response in mild NCD which can be further evaluated in a larger scale RCT.

There are also limitations associated with this pilot trial. Due to the small sample size, the study findings may have limited generalizability and produce imprecise and potentially inflated effect size estimates for any statistically significant findings. Therefore, the power calculation for the main RCT will be based on the minimally clinically important difference (MCID) rather than the observed pilot effect sizes (36). The absence of a control group is another drawback associated with this study and may lead to a high risk of bias. Lastly, as a single-site study and limiting participation to English-speaking individuals, both will restrict the generalizability of the results, introduce bias and not represent a diverse global population.

## Conclusion

Mild NCD is becoming increasingly prevalent and poses a significant global health care concern due to the lack of definitive treatment options. Non-invasive neuromodulation techniques such as dTMS may prove to be a promising treatment possibility for mild NCD. This pilot study will provide preliminary data pertaining to the tolerance and acceptability of dTMS treatment, and secondarily, its impact on cognition in individuals with mild NCD. Additionally, it will also provide insights into the blood and electrophysiological biomarkers associated with mild NCD. Collectively, findings from this study will provide foundational groundwork to rigorously evaluate dTMS as a novel treatment intervention for mild NCD through a full-scale RCT.

## Data Availability

No datasets were generated or analysed during the current study. All relevant data from this study will be made available upon study completion.

## CRedIT Authorship Contribution

**Mehak Ismail Rajani:** Conceptualization, Investigation, Methodology, Project Administration, Visualization, Writing – original draft,

**Horodjei Yaya:** Conceptualization, Investigation, Project Administration, Resources, Writing – Review & Editing

**Emily Vandehei**: Conceptualization, Investigation, Project Administration, Resources, Writing – Review & Editing

**Anne-Marie Di Passa:** Conceptualization, Project Administration, Writing – Review & Editing

**Carly McIntyre-Wood**: Conceptualization, Investigation, Writing – Review & Editing

**Shelby Prokop-Millar**: Conceptualization, Writing – Review & Editing

**Daniel Krzyzanowski**: Resources, Writing – Review & Editing

**Molly Zhang**: Investigation, Resources, Writing – Review & Editing

**Allan Fein**: Conceptualization, Resources, Writing – Review & Editing

**Emily MacKillop:** Conceptualization, Supervision, Writing – Review & Editing

**Jane De Jesus:** Resources, Writing – Review & Editing

**Benicio Frey:** Writing – Review & Editing

**James MacKillop:** Conceptualization, Methodology, Supervision, Funding Acquisition, Writing – Review & Editing

**Dante Duarte:** Conceptualization, Methodology, Supervision, Funding Acquisition, Writing – Review & Editing

All authors commented on previous versions of the manuscript. All authors read and approved the final manuscript.

